# Detection, prevalence, and duration of humoral responses to SARS-CoV-2 under conditions of limited population exposure

**DOI:** 10.1101/2020.08.14.20174490

**Authors:** Tyler J. Ripperger, Jennifer L. Uhrlaub, Makiko Watanabe, Rachel Wong, Yvonne Castaneda, Hannah A. Pizzato, Mallory R. Thompson, Christine Bradshaw, Craig C. Weinkauf, Christian Bime, Heidi L. Erickson, Kenneth Knox, Billie Bixby, Sairam Parthasarathy, Sachin Chaudhary, Bhupinder Natt, Elaine Cristan, Tammer El Aini, Franz Rischard, Janet Campion, Madhav Chopra, Michael Insel, Afshin Sam, James L. Knepler, Andrew P. Capaldi, Catherine M. Spier, Michael D. Dake, Taylor Edwards, Matthew E. Kaplan, Serena Jain Scott, Cameron Hypes, Jarrod Mosier, David T. Harris, Bonnie J. LaFleur, Ryan Sprissler, Janko Nikolich-Žugich, Deepta Bhattacharya

**Author notes:** These authors contributed equally and are listed in alphabetical order (Tyler J. Ripperger, Jennifer L. Uhrlaub, Makiko Watanabe, Rachel Wong). these authors contributed equally. Address correspondence to Janko Nikolich-Žugich, at; or Deepta Bhattacharya, at, or by mail at P.O. Box 249221, Department of Immunobiology, University of Arizona College of Medicine-Tucson, 1501 N. Campbell Ave., Tucson, AZ 85724.

## Abstract

We conducted an extensive serological study to quantify population-level exposure and define correlates of immunity against SARS-CoV-2. We found that relative to mild COVID-19 cases, individuals with severe disease exhibited elevated authentic virus-neutralizing titers and antibody levels against nucleocapsid (N) and the receptor binding domain (RBD) and the S2 region of spike protein. Unlike disease severity, age and sex played lesser roles in serological responses. All cases, including asymptomatic individuals, seroconverted by 2 weeks post-PCR confirmation. RBD- and S2-specific and neutralizing antibody titers remained elevated and stable for at least 2-3 months post-onset, whereas those against N were more variable with rapid declines in many samples. Testing of 5882 self-recruited members of the local community demonstrated that 1.24% of individuals showed antibody reactivity to RBD. However, 18% (13/73) of these putative seropositive samples failed to neutralize authentic SARS-CoV-2 virus. Each of the neutralizing, but only 1 of the non-neutralizing samples, also displayed potent reactivity to S2. Thus, inclusion of multiple independent assays markedly improved the accuracy of antibody tests in low seroprevalence communities and revealed differences in antibody kinetics depending on the viral antigen. In contrast to other reports, we conclude that immunity is durable for at least several months after SARS-CoV-2 infection.

## INTRODUCTION

SARS-CoV-2, the causative agent of COVID-19, has infected over 20 million people worldwide, with over 750,000 dead as of August 13, 2020. Serological testing for SARS-CoV-2 antibodies is an important tool for measuring individual exposures, community transmission, and the efficacy of epidemiological countermeasures. While a few epicenters of infection have seen relatively robust spread of the virus (Rosenberg et al., 2020; Stadlbauer et al., 2020), COVID-19 prevalence in most of the world has been low. For example, studies in Spain and Switzerland revealed overall seroprevalences of ~5%, with some communities at just 1% antibody positivity (Pollán et al., 2020; Stringhini et al., 2020). The challenges of accurate antibody testing for SARS-CoV-2 in low seroprevalence communities have led to several unexpected conclusions. As an example, a seroprevalence study in Santa Clara county, California suggested higher infection rates than had been anticipated, thereby leading to the interpretation that SARS-CoV-2 was much less deadly than originally thought (Bendavid et al., 2020). Yet this conclusion was problematic given that the false positive rates of the administered test approached the true seroprevalence of the community (Bennett and Steyvers, 2020). Thus, it is likely that many positive results were inaccurate, and the overall infection fatality rate was substantially higher than estimated in this study (Bennett and Steyvers, 2020). Reducing this false positive rate is critical for accurate seroprevalence studies. Moreover, serological testing has an additional imperative to guard against false positive results that could entice the subject to falsely assume immunity where none may exist. Indeed, the assumption of immunity associated with a positive test result may be amongst the primary motivations for participation in these serological surveys. Virus neutralization assays are functional correlates of immunity but require Biosafety Level 3 facilities and are difficult to scale and deploy as clinical assays. Yet tests that fail to provide confidence in functional immune status undermine this important epidemiological tool. Finally, poor positive predictive values are especially problematic in the context of convalescent plasma donations, where most samples would be ineffective in passive transfer therapies.

Serological studies have also been used to estimate the durability of antibody production and immunity after SARS-CoV-2 infections. Here again, several surprising conclusions have been reached regarding the short duration of immunity, with several studies suggesting that in a substantial number of subjects, antibody levels wane to below the limit of detection within a matter of weeks to months (Ibarrondo et al., 2020; Long et al., 2020a; Pollán et al., 2020; Seow et al., 2020). Yet all T-dependent humoral responses, even ones that are exceptionally durable, begin with an initial wave of short-lived plasma cells which decline quickly and are progressively replaced by a smaller number of longer-lived antibody-secreting plasma cells (Amanna, 2007; Manz et al., 1997; Slifka et al., 1998; Sze et al., 2000). Thus, the decay in antibody production after infection or vaccination is not linear and cannot be extrapolated from early timepoints, demonstrating the need for longer-term follow-up studies. Indeed, such short-term antibody production would be without precedent following acute coronavirus infections, which typically induce immunity for at least a year and for SARS-CoV-1, often for much longer (Callow et al., 1990; Guo et al., 2020; Reed, 1984; Tan et al., 2020). Keys to the accurate interpretation of such studies are sensitive assays, PCR confirmation of test cases, and longitudinal tests of seropositive individuals. Authentic virus neutralization assays are also useful as true correlates of immunity (Zinkernagel and Hengartner, 2006). Absent these components, conclusions about the duration of immunity are premature.

Here, we successfully employed a strategy using RBD and S2 as antigenically distinct tests to accurately identify seropositive individuals in the community. In doing so, this assay greatly reduced the existing limitations to testing accuracy in low seroprevalence communities and identified individuals for subsequent analysis of the immune response. We found that disease severity, but not age or sex, were correlates of the magnitude of the response. Further, use of these two antigens, nucleocapsid protein, and neutralizing antibody titers revealed discordance in the durability of antibody responses depending on the viral protein. In contrast to earlier reports, we demonstrate durable production of functionally important antibodies lasting at least 2-3 months post-disease onset.

## RESULTS

Numerous serological tests that have received Food and Drug Administration Emergency Use Authorizations (https://www.fda.gov/medical-devices/coronavirus-disease-2019-covid-19-emergency-use-authorizations-medical-devices/eua-authorized-serology-test-performance) rely on reactivity to the SARS-CoV-2 RBD domain of the S protein (Amanat et al., 2020; Premkumar et al., 2020). To begin validation of a serological assay for antibodies to RBD, we tested 43 serum samples from PCR-confirmed COVID-19 patients in the hospital at various stages of disease, 48 convalescent samples, and 23 samples from healthy donors. Serum dilution ELISAs were performed to quantify RBD-reactive antibodies in these samples. Mammalian RBD antigen preparations were selected as targets, as they demonstrated superior signal:noise ratios relative to bacterially-produced protein (**Figure S1A**). Antibody titers were quantified as area under the curve (AUC) and correlated with neutralization of the live USA- WA1/2020 strain of SARS-CoV-2, rather than S-protein pseudotyped virus (Giroglou et al., 2004), due to the poor agreement between these functional assays (**Figure S1B**), and because of the modest sensitivity of some pseudovirus neutralization tests relative to those of authentic virus (Schmidt et al., 2020). The correlation was strong between RBD-reactive IgG and plaque reduction neutralization test (PRNT) titers, which we quantified as the final dilution at which 90% viral neutralization occurred (PRNT_90_) (**Figure 1A**). RBD-reactive IgM antibodies also correlated with PRNT_90_ titers (**Figure S1C**). Because (i) IgM and IgG ratios are not indicative of the timing of disease onset (Hou et al., 2020; Long et al., 2020b; Qu et al., 2020), (ii) IgA is induced by SARS-CoV-2 (Isho et al., 2020; Iyer et al., 2020; Sterlin et al., 2020), and (iii) both IgG and IgM isotypes correlated with neutralizing titers, we chose to quantify total (all isotypes) antigen-specific antibodies for seroprevalence studies.

**Figure 1:**
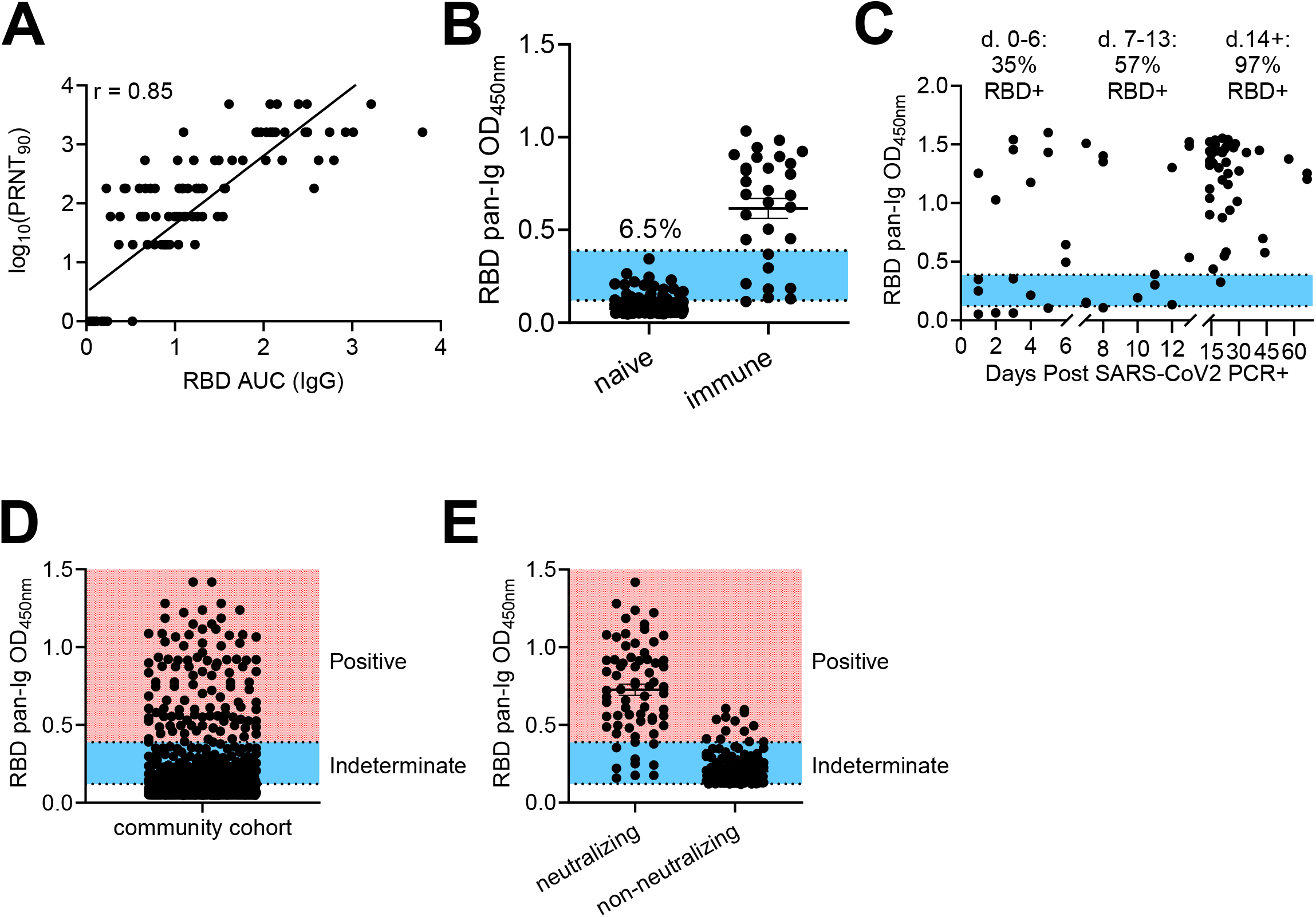
Assessment of RBD-based sensitivity and specificity in serological testing. **(A)** Serum samples from healthy controls and confirmed COVID-19 cases were assessed for RBD reactivity by ELISA and neutralization of live SARS-CoV-2. PRNT_90_ values were determined as the last dilution by which 90% neutralization occurred. Antibody titers were quantified for RBD by quantifying area under the curve (AUC) across a serial dilution curve. r values were calculated by Pearson’s Correlation Test. (**B**) Pre-2020 negative control samples (352) and 30 samples from SARS-CoV-2 exposed individuals were screened by ELISA at a single 1:40 dilution against RBD. The blue region indicates overlap of OD values between negative and positive control samples. % indicates frequency of negative control values in this range. Experiments were repeated 3 times. (**C**) RBD seroreactivity was quantified based upon time elapsed from PCR+ confirmation of SARS-CoV-2 infection. (**D**) Individuals recruited from the community (5882) were screened for seroreactivity to RBD. (**E**) PRNT_90_ analysis from community drawn samples that displayed indeterminate or positive RBD seroreactivity. Samples that neutralized 90% of virions at least at a 1:20 dilution were considered positive. Experiments were repeated at least once.

To determine if RBD was capable of distinguishing between SARS-CoV-2 exposed and uninfected individuals and to set preliminary thresholds for positive calls, we initially tested 1:40 serum dilutions of samples from 30 PCR+ SARS-CoV-2 infected individuals and 32 samples collected prior to September, 2019, well before the onset of the current pandemic (**Figure S1D**). Using this test data set, we established a preliminary positive cutoff OD_450_ value of 0.12, equal to 3 standard deviations above the mean values of the negative controls. We next used this preliminary threshold to test an expanded cohort of 320 negative control samples collected prior to 2020. **(Figure 1B)**. Reactivity to RBD was clearly distinguishable for the majority of positive samples from negative controls (**Figure 1B**). However, 6.5% of the expanded negative control group displayed RBD reactivity that overlapped with PCR+ individuals (**Figure 1B, blue shade**), some of whom may have been early into disease and had not yet generated high levels of antibodies. To quantify the sensitivity of the assay relative to time of diagnosis, we measured antibody levels to RBD and plotted these values against time following SARS-CoV-2 PCR+ confirmation. Whereas the sensitivity was modest within the first two weeks, after 2 weeks, 42 of 43 samples showed high ELISA signal (**Figure 1C)**. Based on these data, samples were considered seropositive at OD_450_ numbers above 0.39, a value slightly above the highest OD obtained from the 352 subjects in the negative control group (**Figure 1B**). Sera were considered negative at OD_450_ values below 0.12. Finally, we created an indeterminate call at OD_450_ values between 0.12-0.39, as we observed some overlap between negative controls and PCR-confirmed samples in this range (**Figure 1B, blue shade**).

We next applied this assay to community testing and obtained serum samples from 5882 self-recruited volunteers from Pima County. Donors included healthcare workers (~26%), first responders (~27%), University of Arizona students (~5%), and other members of the general public (~42%). Currently febrile or otherwise symptomatic patients were excluded. Sera from 73 individuals preliminarily scored as seropositive (**Figure 1D**). These samples, along with another 171 samples with OD_450_ values in the indeterminate range were tested for virus neutralization at a serum dilution of 1:20 (**Figure 1E**). Nine samples with RBD OD_450_ values below 0.39 were observed to neutralize SARS-CoV-2 (**Figure 1E**). More problematically, we found that 13 of the 73 samples (17.8%) called positive by RBD-reactivity failed to neutralize authentic SARS-CoV-2 (**Figure 1E**). If virus neutralization is considered as a measure of ‘true’ seropositivity, RBD ELISAs alone provided a relatively modest positive predictive value of 82%. These observations indicated a clear need for a secondary screen to accurately quantify seropositivity in a community with low infection rates.

To improve the positive predictive value, we considered the use of an orthogonal antigenically distinct test. We first tested nucleocapsid (N) protein, as several other commercial serological tests quantify antibodies to this antigen (Bryan et al., 2020; Burbelo et al., 2020). IgG antibody titers to N protein in our collected sample cohort showed a strong correlation to PRNT_90_ titers (**Figure 2A**). A weaker correlation was observed between N-reactive IgM levels and PRNT_90_ titers (**Figure S2A**). We next assayed reactivity to N antigen using a subset of the pre-2019 validation samples employed for RBD. N protein seroreactivity overlapped substantially between negative and positive controls (**Figure 2B**). Moreover, 5 confirmed COVID-19 samples showed very weak reactivity to N (**Figure 2B**). Because of the relatively poor performance of N protein as an antigen in our hands, we next tested the S2 domain of S protein as another candidate to determine seropositivity. RBD is located on the S1 domain, rendering S2 antigenically distinct (Bosch et al., 2003; Li, 2016; Wrapp et al., 2020). IgG antibody titers to S2 correlated well with PRNT_90_ titers (**Figure 2C)**. Assessment of S2 serum reactivity in the pre-2019 cohort revealed that approximately 3.3% of these samples overlapped with signals in PCR-confirmed COVID-19 samples (**Figure 2D)**. We thereafter employed a threshold of OD_450_>0.35, as our cutoff for S2 positivity, which was 5 standard deviations above the average seroreactivity from the original 32-samples from the negative control cohort. Specificity control testing using 272 negative control sera showed that reactivities of negative samples against RBD and S2 were largely independent of one another, as samples with high signal for one antigen rarely showed similar background for the other (**Figure 2E**). Based on these data, we chose to rely on combined RBD and S2-reactivities as accurate indicators of prior SARS-CoV-2 exposure.

**Figure 2:**
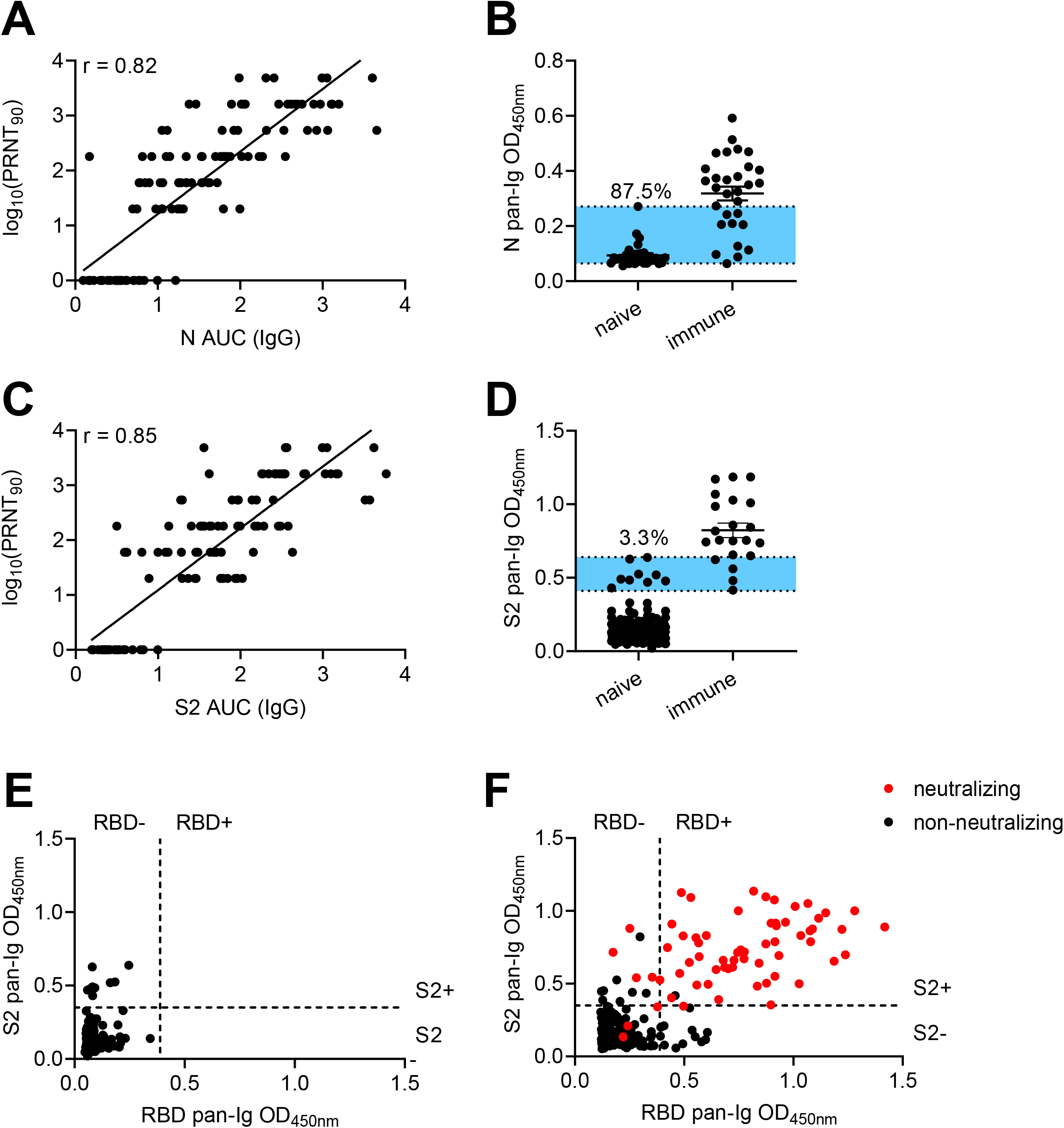
Assessment of S2 and N antibodies as secondary confirmations of seropositivity. (**A**) Correlations of neutralization and N-specific IgG ELISA titers across 115 serum samples from healthy controls and COVID-19 cases. (**B**) A sample set of 32 pre-pandemic controls and 30 PCR+ SARS-CoV-2 samples were assayed for seroreactivity to N protein. Blue shaded region indicates overlap between negative and positive controls. Frequency of negative controls in this range is shown. (**C**) Correlations of neutralization and S2-specific IgG ELISA titers across 114 serum samples from healthy controls and COVID-19 cases. (**D**) Pre-pandemic negative control samples (272) were screened for seroreactivity against S2 and compared to 30 PCR-confirmed SARS-CoV-2-exposed sera. (**E**) Comparison of RBD and S2 seroreactivity across 272 pre-pandemic serum samples. (**F**) ELISA results from indeterminate and putative seropositive samples from community testing. Thresholds for seropositivity were defined as in (**E**). Red circles indicate samples that have PRNT_90_ titers of at least 1:20. Experiments were repeated at least once.

With this improved combinatorial RBD and S2 assay to exclude false positives, we re-examined the original samples from the cohort of 5882 subjects that displayed RBD OD_450_ values greater than 0.12 (**Figure 1D-E**). Of the 13 non-neutralizing samples that displayed high (OD_450_ >0.39) RBD reactivity, 12 lacked S2 reactivity (**Figure 2F**). In contrast, the remaining 60 RBD+ neutralizing samples all displayed substantial reactivity to S2 (**Figure 2F**). Five of the 9 samples that fell below the RBD cutoff, yet still neutralized virus, displayed strong reactivity to S2 (**Figure 2F**). Based on these data, we established a scoring criterion of RBD OD_450_>0.39, S2 OD_450_>0.35 as seropositive; RBD OD_450_ between 0.12-0.39, S2 OD_450_>0.35 as indeterminate; and all other samples as seronegative. Applying these criteria to 320 samples obtained prior to 2020 would lead to 317 negative, 3 indeterminate, and 0 positive calls. Using these same criteria, we achieved an empirically defined false positive rate of just 0.02%, with only 1 positive sample incapable of neutralizing live SARS-CoV-2 virus. Approximately half the samples called as indeterminate contained neutralizing antibodies. Only 3 samples called as negative possessed neutralizing titers, which were usually low (1:20). To further confirm the sensitivity of the assay, we tested 993 samples at random for neutralizing antibodies. Of these, none of the samples called as negative possessed neutralizing activity (data not shown). These data demonstrate that inclusion of S2 as a requisite confirmatory screen markedly improves the positive predictive value of SARS-CoV-2 serological assays, especially in areas with low SARS-CoV-2 seroprevalence.

Several recent reports have suggested more robust immune responses in those with severe disease relative to mild cases (Choe et al.; Ko et al., 2020; Long et al., 2020a; Qu et al., 2020). Moreover, the ratios of S and N antibody specificities correlate with disease outcome (Atyeo et al., 2020). We therefore examined our data for these trends. First, in our PCR-confirmed cohort, we plotted IgG titers relative to the time of disease onset, stratified by disease severity. Severe disease (hospital admission) correlated with significantly higher antibody titers against RBD, S2, and N than those with mild disease, who were symptomatic but did not require hospital admission (**Figure 3A-C**). Neutralizing titers were also higher in those with severe disease relative to mild cases (**Figure 3D**). Through campus screening efforts, we also identified 6 PCR+ individuals who either never developed symptoms or had only a brief and mild headache or anosmia. Although previous reports suggested that such individuals may infrequently seroconvert or frequently serorevert (Long et al., 2020a; Sekine et al., 2020), all such individuals in our cohort showed seroreactivity to RBD, S2, and all but one to N (**Figures S3A-C**), consistent with other recent studies (Choe et al.; Ko et al., 2020). Given that older adults, as well as those of male sex, exhibit disproportional morbidity and mortality from COVID-19, we also sought to test whether humoral immunity in these subjects may be quantitatively reduced (Liu et al., 2020). Contrary to this expectation, we did not observe any adverse impact of advanced age on humoral immunity (**Figure 3E-H**). Similarly, within our cohort, females and males had similar anti-RBD, N, S2, and neutralizing responses (**Figure S3D-G)**.

**Figure 3:**
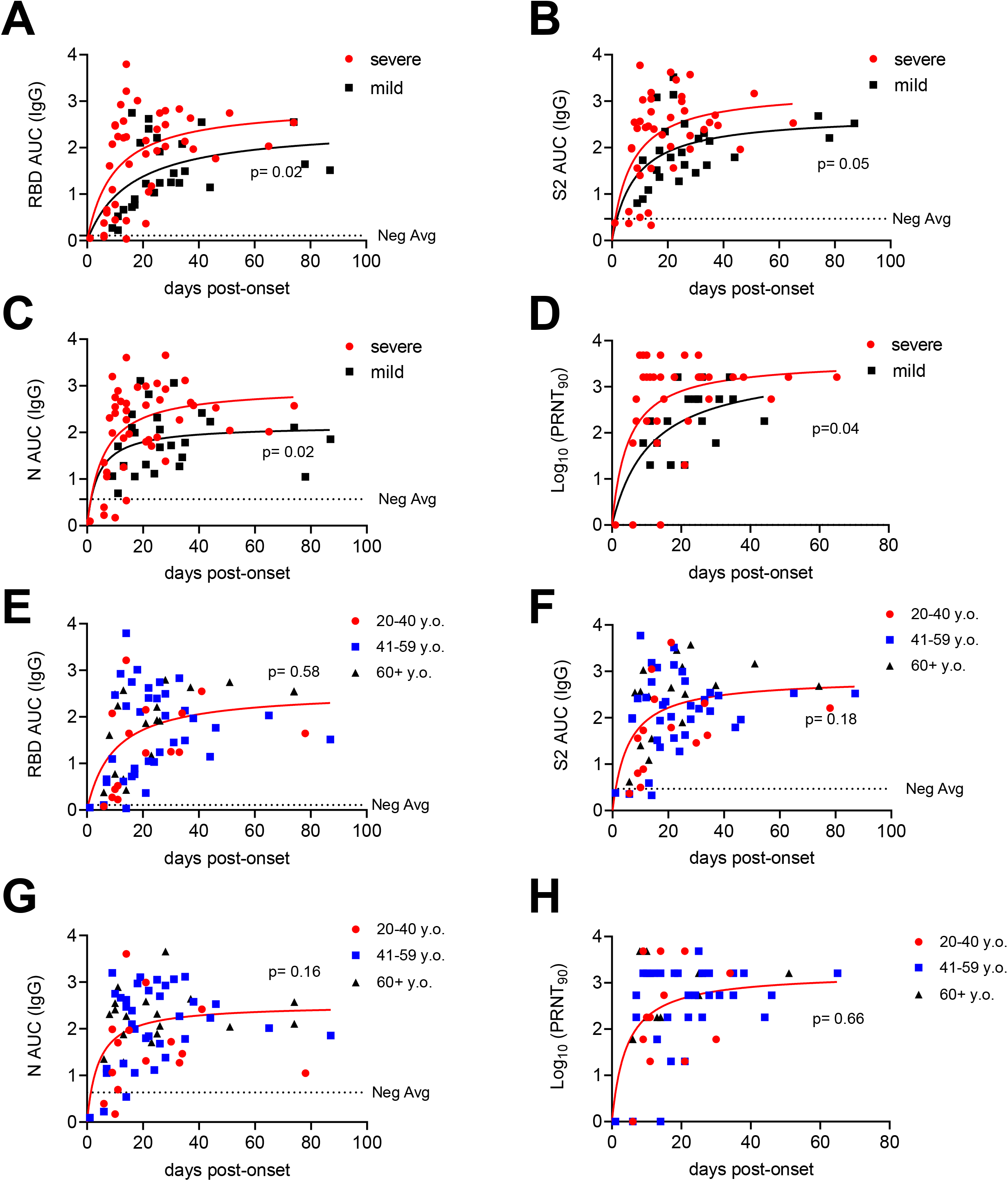
Antibody responses to SARS-Cov2 as a function of disease severity and age. **(A-C)** Antibody titers to RBD (A), S2 (B), and N (C), over time post-onset of SARS-CoV-2 infection symptom grouped by case severity. The negative control average was determined by calculating the average AUC value of negative control (n=25) samples. P values represent comparison of fit in non-linear regression model between displayed groups. **(D)** PRNT_90_ values over time post-onset of SARS-CoV-2 infection symptoms. P values were calculated as in (A). **(E-H)** Antibody titers over time post-onset of SARS-CoV-2 infection symptoms from PCR+ confirmed patients or seropositive individuals from community wide cohort for RBD (E), N (F), and S2 (G), grouped by patient age. **(H)** PRNT_90_ values over time post-onset of SARS-CoV-2 infection symptoms grouped by patient age.

Individuals with mild disease have been reported to lose SARS-CoV-2-specific antibodies quickly into convalescence (Ibarrondo et al., 2020; Long et al., 2020a; Seow et al., 2020). To assess the durability of antibody production in our cohort, we first returned to the community cohort of 5882 individuals. Twenty-nine of the seropositive subjects had reported mild symptoms consistent with COVID-19. These positive samples were thus plotted alongside PCR-confirmed mild disease cases against time post-disease onset to determine if any trends could be observed in declining antibody levels. Across subjects, IgG specific for RBD (**Figure 4A**) and S2 (**Figure 4B**) appeared to peak near 30 days post-onset and then partially decline before settling to a more stable nadir at later timepoints, as would be expected for all acute viral infections. We considered the possibility that we may have missed subjects that had seroreverted prior to their antibody test, thereby incorrectly raising our estimates of the durability of antibody production. Therefore, to examine the duration of IgG production in more depth, a subset of seropositive individuals with relatively low titers was tested longitudinally up to 122 days post-onset. These data again revealed stable RBD and S2 IgG levels at later stages of convalescence (**Figures 4A-B**). However, N-reactive IgG levels were quite variable and approached the lower limit of detection in several subjects at later timepoints (**Figure 4C**). A direct comparison in matched subjects of the changes in RBD, S2, and N IgG titers over time confirmed the variability in N responses and rapid decline in a subset of individuals (**Figure 4D**). Most importantly, neutralizing antibody levels remained high with very little decay as a function of time (**Figure 4E**). These data suggest stable neutralizing, RBD, and S2-specific antibodies, but variable and often declining N-reactive titers during convalescence. Together, these data are consistent with the maintenance of functionally important antibody production for at least several months after infection, and caution against the use of α-N antibodies to estimate immunity or seroprevalence.

**Figure 4:**
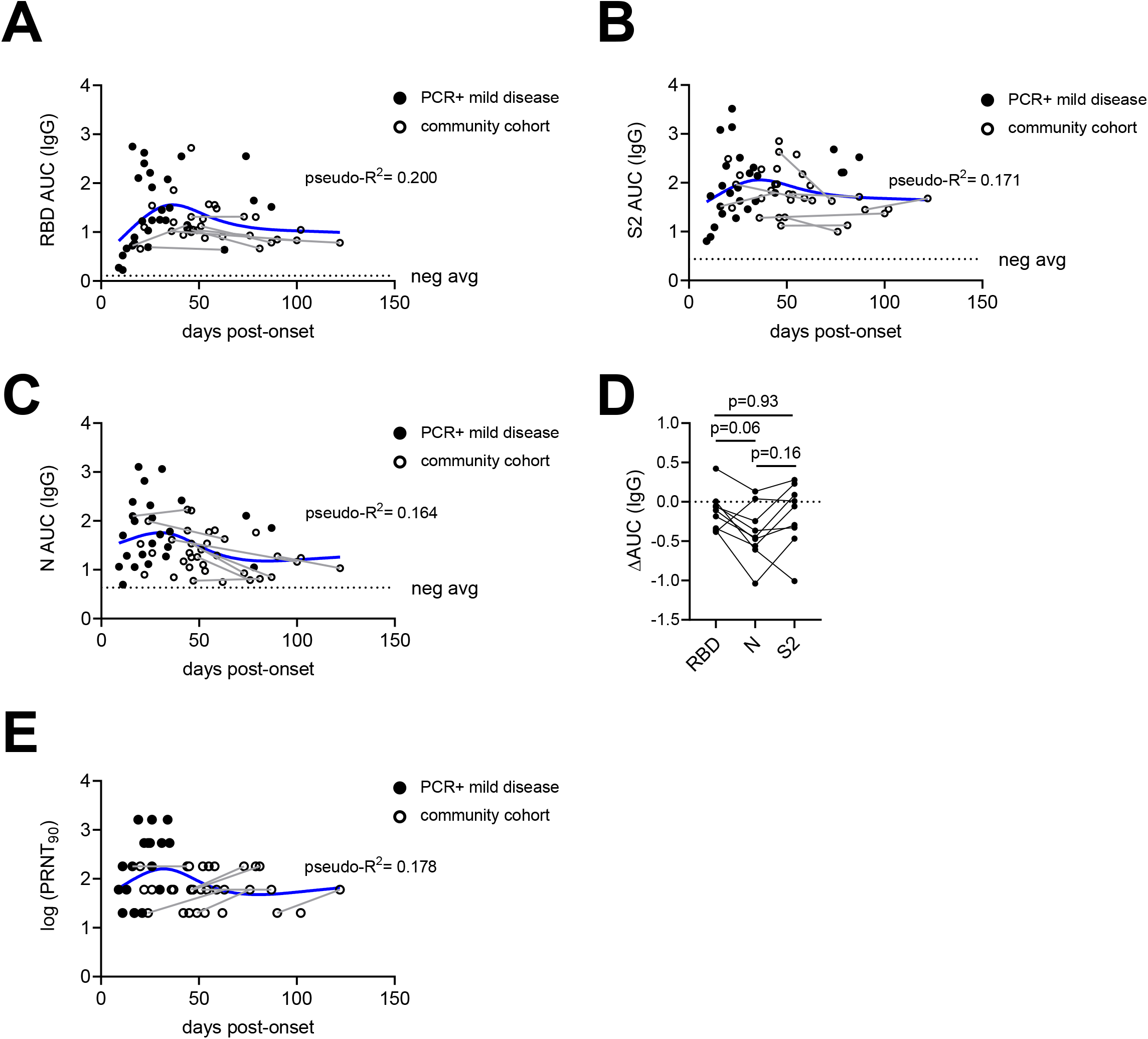
Antibody Responses to Spike Glycoprotein are more stable than responses to Nucleocapsid. **(A-C)** Antibody titers for mild infections over time to RBD (A), S2 (B), and N (C) for PCR-confirmed subjects and seropositive samples from community serological testing. Solid lines connect data from individuals sampled serially over time. Blue line depicts smoothing splines curve fit with 4 knots. Dashed line depicts mean values from seronegative controls. **(D)** Subjects sampled serially were assessed for changes in antibody titers to RBD, S2, and N from the first draw to the last draw collected. Only subjects in which the last draw occurred >6 weeks from onset are shown. P values were calculated by paired 1-way ANOVA. **(E)** Neutralizing titers were measured for longitudinal subjects over time post-onset. Solid lines connect data from individuals sampled serially over time. Curve (blue line) was generated in using smoothing splines with 4 knots.

## DISCUSSION

Here, we demonstrated that using two antigenically distinct serological tests can greatly remedy specificity problems that are exacerbated in low SARS-CoV-2 seroprevalence communities. RBD and S2 seroreactivity behaved independently for SARS-CoV-2-unexposed individuals, thereby suggesting that the theoretical false positive rate of the overall assay is the product of the two tests. Using neutralization assays to confirm these results, we found our empirically determined false positive rate to be <0.02% (1/5882), consistent with the independence of the RBD and S2 tests. The tight co-incidence between RBD/S2 positivity and the presence of neutralizing antibodies, even in low seroprevalence populations, is especially valuable for identifying individuals who likely have some degree of immunity and could potentially serve as convalescent plasma donors. Surprisingly, nucleocapsid (N), which is used by several commercial serological tests as an antigen, did not perform as well in our assays, with high false positive and negative rates.

Though we are uncertain why N protein reactivity proved less discriminatory in our hands relative to published work (Bryan et al., 2020; Steensels et al., 2020), as one possible explanation, we observed that in several subjects, N-specific antibodies declined more rapidly than those against RBD or S2. This unexpected finding may in part help explain some discrepancies in the literature. In some reports, SARS-CoV-2-specific N antibodies fell to undetectable levels within 2-3 months in up to 40% of those recovering from mild disease (Long et al., 2020a; Pollán et al., 2020), which would be remarkably transient and very unusual for acute viral infections, even other common coronaviruses (Callow et al., 1990; Reed, 1984). Although most N titers did not fall fully below our detection limits, we also observed such a decline in some subjects. Yet encouragingly, neutralizing antibodies and those against RBD and S2 reached a stable nadir after the initial expected decline, presumably as short-lived plasma cells were replaced with long-lived antibody secreting cells. These data are consistent with expectations for acute viral infections and with the conclusions of other studies currently on preprint servers (Isho et al., 2020; Iyer et al., 2020; Wajnberg et al., 2020). In this regard, the primary data for S and neutralizing antibody responses seem consistent across several studies (Ibarrondo et al., 2020; Seow et al., 2020), though the interpretations differ. These differences in interpretation are reminiscent of studies on the length of SARS-CoV-1 immunity. Early reports suggested that immunity was transient (Cao et al., 2007), but more recent studies have demonstrated that SARS-CoV-1 neutralizing antibodies can still be detected 12-17 years afterwards (Guo et al., 2020; Tan et al., 2020). Given these lessons, conclusions about the rapid loss of immunity to SARS-CoV-2 are premature and inconsistent with the data we presented here.

The reasons for the differences in antibody responses across antigens are difficult to explain, given the identical inflammatory environment in which these responses arose. One possibility is that the avidities of germline precursors differ for N- and S-protein specificities. For both memory and plasma cells, there appears to be a ‘sweet spot’ of antigen avidity that promotes optimal responses (Abbott et al., 2018). A second possibility is that N-protein responses are driven by cross-reactive memory, rather than naïve B cells. Memory B cells are substantially more diverse than are plasma cells, thereby encoding a hidden repertoire that is not represented in serum antibodies (Lavinder et al., 2014; Purtha et al., 2011; Smith et al., 1997). Consistent with this possibility, N protein is more conserved across coronaviruses than is RBD (Srinivasan et al., 2020). Memory responses, especially by isotype-switched B cells, are directed by fundamentally distinct transcriptional programs than those of naïve cells (Bhattacharya et al., 2007; Jash et al., 2016; Wang et al., 2012; Zuccarino-Catania et al., 2014). For example, the transcription factor ZBTB32 specifically limits the magnitude and duration of memory B cell responses, perhaps to keep chronic infections from overwhelming the system (Jash et al., 2016, 2019). It remains to be established whether such mechanisms may be selectively operating on SARS-CoV-2 and other coronavirus N antibody responses due to their antigenic similarity between strains.

Taken together, we have reported a highly specific serological assay for SARS-CoV-2 exposure that is usable in very low seroprevalence communities, and that returns positive results that are highly co-incident with virus neutralization. Using this assay, we characterize the responses in different subject populations by age, sex and disease severity, we demonstrate that antibody production persists for at least 3 months, and we suggest explanations for some reports that concluded otherwise.

### Limitations of current study

The above assay allowed us to examine the influence of age, sex, and disease severity on levels of humoral immunity in our tested populations. Similar to other studies (Qu et al., 2020; To et al., 2020), we found that severe disease correlated positively with levels of antibody immunity. While both older adults (>50 and even more >65 years of age) and males are more vulnerable to COVID-19 (Klein et al., 2020a, 2020b; Palaiodimos et al., 2020), levels of humoral immunity did not reveal age or sex-related differences that could explain such vulnerability. A caveat here is that our study had a limited longitudinal component and that we could not determine whether there may have been a delay or reduction in humoral immunity at earlier time points of the disease. A second related caveat is that in our community testing cohort we may have missed individuals who were seropositive initially but then seroreverted by the time of the antibody test. Finally, the latest timepoint post-disease onset in our study is 122 days. It remains possible that antibody titers will wane substantially at later times. Additional serial sampling of PCR-confirmed mild cases will be required to test these possibilities.

## Data Availability

Primary data are shown within the manuscript. Additional details are available upon reasonable request.

## ACKNOWLEDGEMENTS

The authors are indebted to the nurses in the intensive care units of Banner University Medical Center – Tucson and South Campuses and research coordinators (Cathleen Wilson and Trina Hughes) for facilitating the collection of samples in critically ill hospitalized patients with COVID-19. We thank F. Krammer, V. Simon, M. Rao, and J. Jhang (Mt. Sinai Hospital) and A. Ellebedy and D. Fremont (Washington University in St. Louis) for reagents and protocols. Supported in part by USPHS awards AG020719 and AG057701 and CDC award 75D30120C08379 (J.N-Ž), the contract CTR050053 from the State of Arizona (J.N-Ž and D.B.), R01AI099108 and R01AI129945 (D.B.), by the COVID-19 Rapid Response Grant from the UArizona BIO5 Institute (to C.C.W., D.B. and J.N-Ž), and the Bowman Endowment in Medical Sciences (J. N-Ž).

## AUTHOR CONTRIBUTIONS

T.J.R., J.L.U., M.W., R.W., R.S., J.N.Z., and D.B. designed the study. T.J.R., J.L.U., M.W., R.W., H.P., C.B., M.K., and R.S. performed experiments. T.J.R., J.L.U., M.W., R.W., A.C., C.S., M.K., T.E., R.S., J.N.Z., and D.B. analyzed the data. T.J.R., J.N.Z., and D.B. wrote the paper. All other authors participated in collection of samples and patient care for the study.

## DECLARATION OF INTERESTS

Unrelated intellectual property of D.B. and Washington University has been licensed by Sana Biotechnology. J.N.Z. is on the scientific advisory board of and receives research funding from Young Blood Inc. R.S. is a founder and chief scientific officer of Geneticure. R.W. is currently an employee of Vir Biotechnology.

**Figure S1 related to Figure 1:**
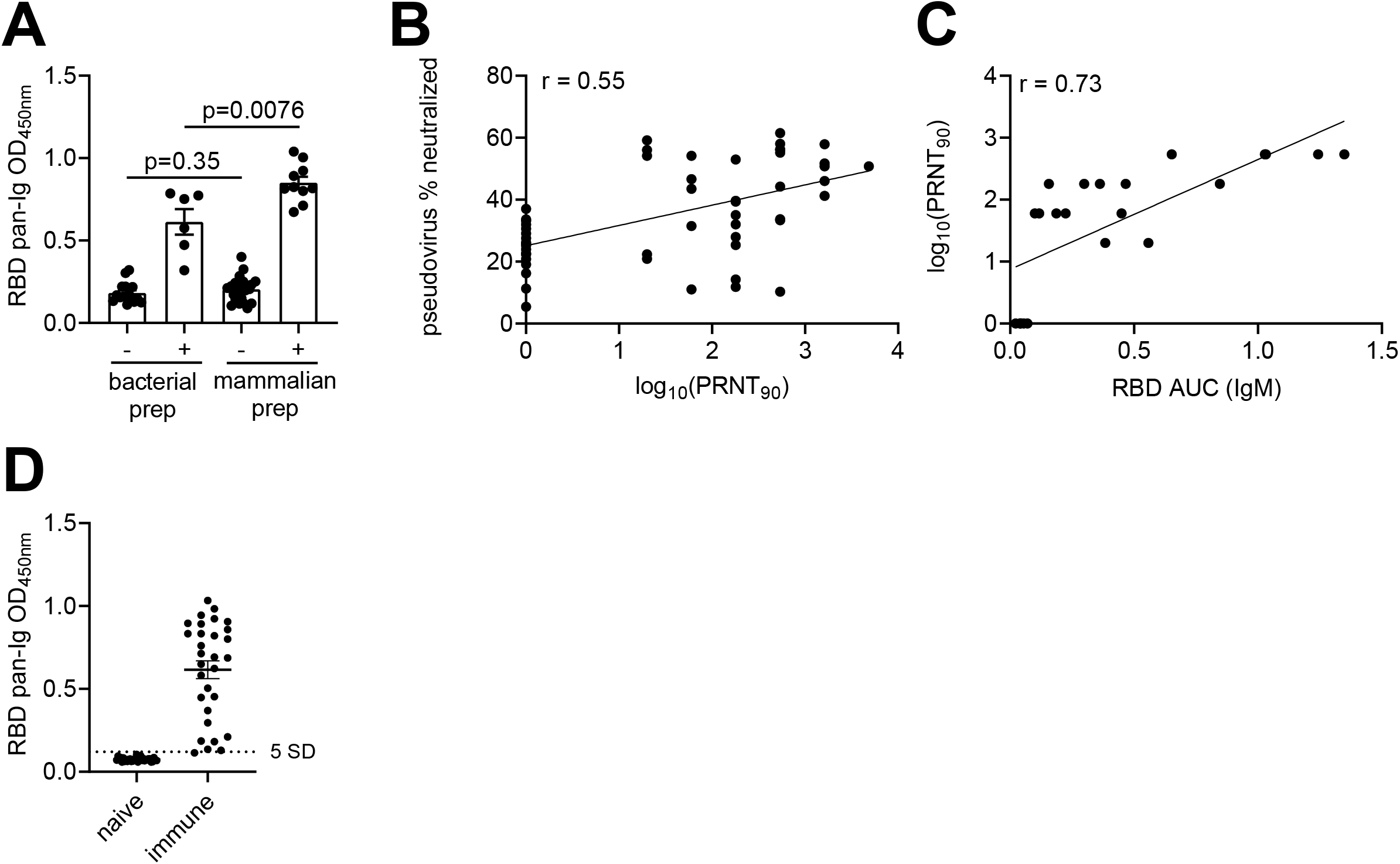
Optimization of RBD-based ELISAs and neutralization assays. (**A**) RBD derived from mammalian and bacterial expression system were compared by ELISA using SARS-CoV-2-neutralizing sera and controls. Statistical differences were determined by students’ unpaired 2-tailed t-test. (**B**) Correlation of pseudovirus neutralization with PRNT_90_ values of live SARS-CoV-2 determined as in Figure 1A. Pseudovirus neutralization titers were determined by % neutralization at a dilution of 1:20. (**C**) Neutralizing titers correlate with IgM antibody titers for RBD. For (B) and (C), r value was determined by Pearson correlation. (**D**) ELISA of pre-2020 sera (32) and COVID-19 samples (30) for RBD reactivity.

**Figure S2 related to Figure 2:**
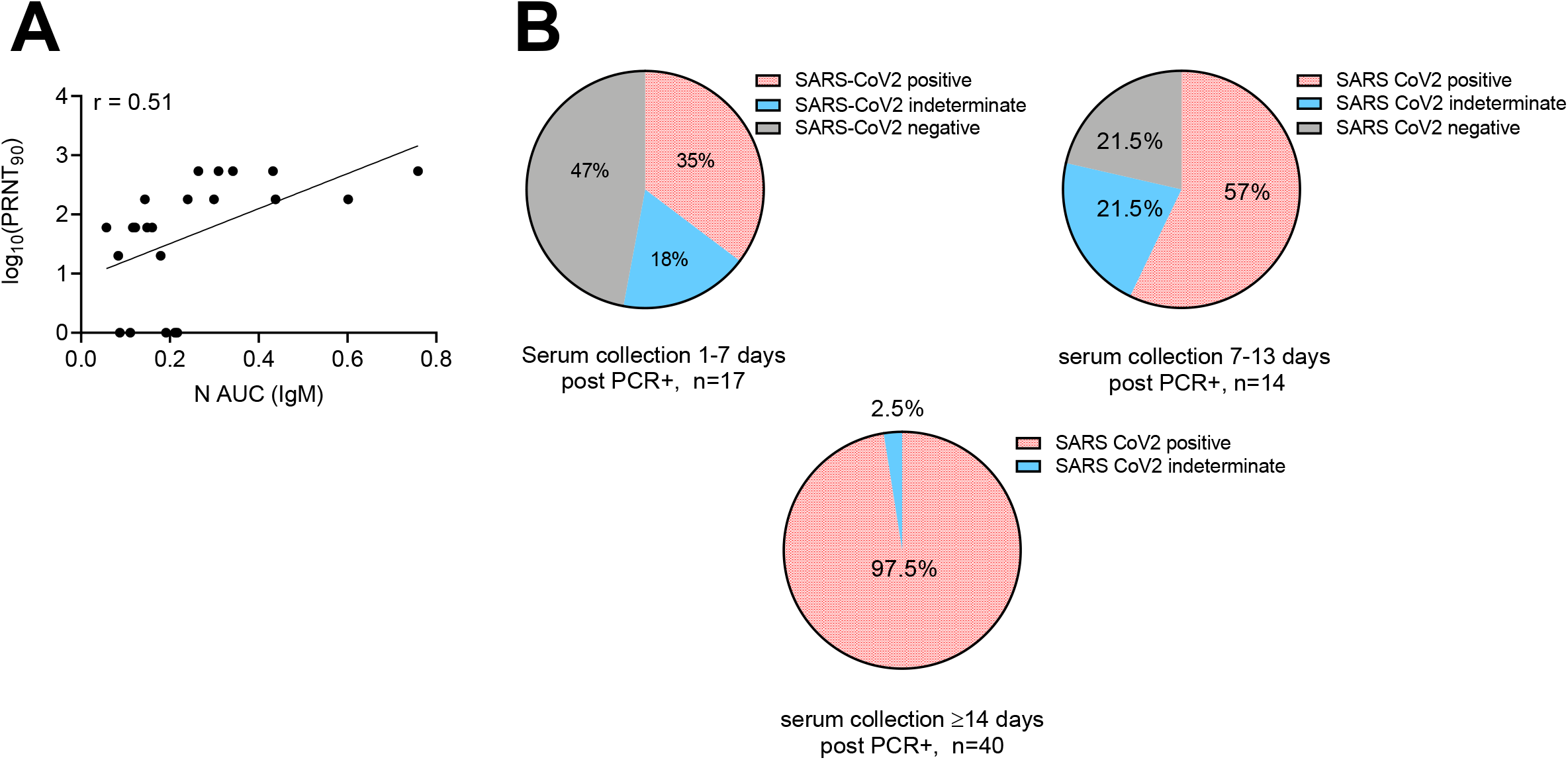
Optimization of secondary screens for S2 and N. (**A**) N-reactive IgM antibody titers were correlated to PRNT_90_ neutralization titers. R value was determined by Pearson’s Correlation. (**B**) Following inclusion of S2 as secondary screen, seropositivity results are shown for individuals collected 1-7 days, 7-13 days, or >14 days post-PCR confirmation. Samples match those used in Figure 1C.

**Figure S3 related to Figure 3:**
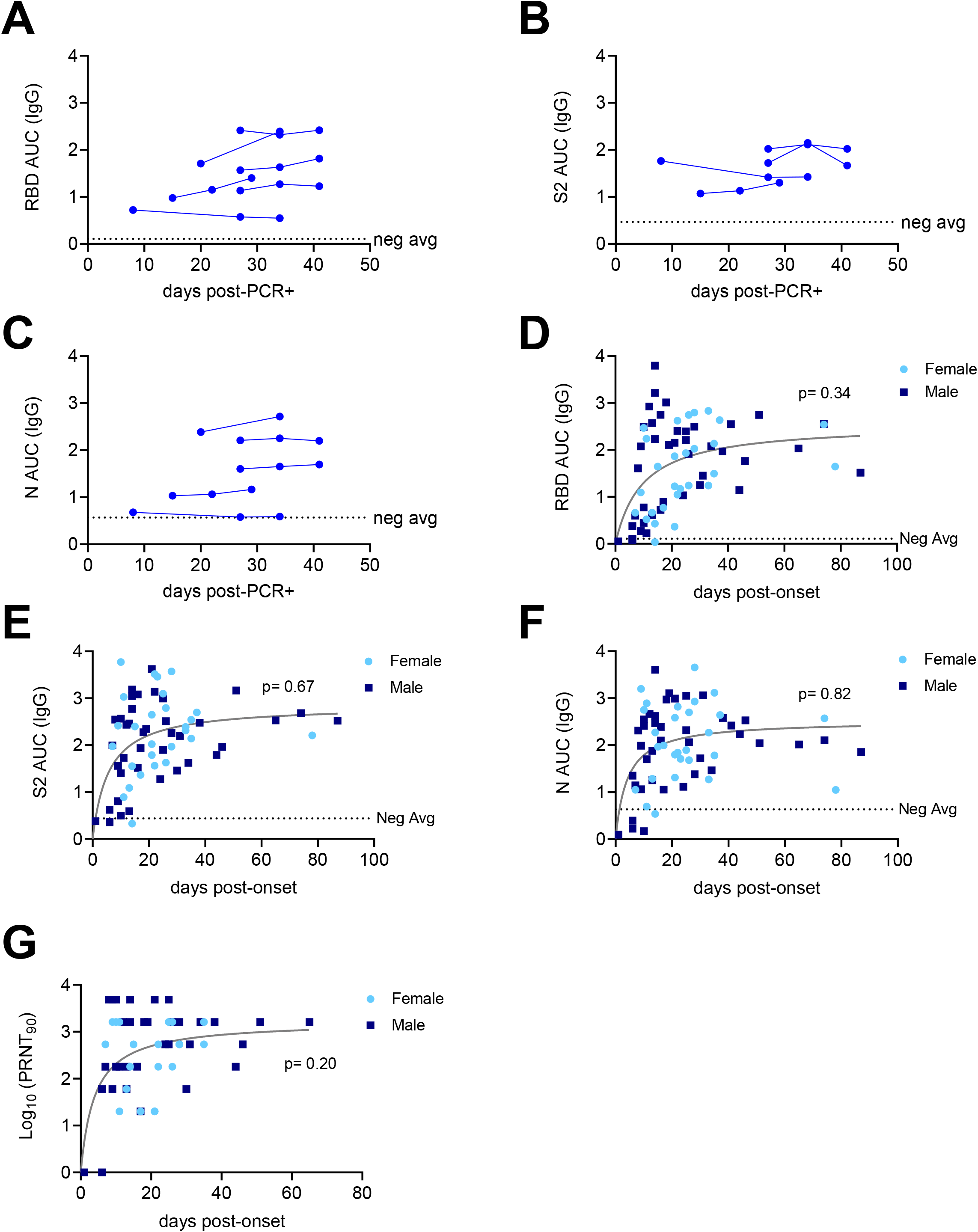
Antibody responses to SARS-Cov2 in asymptomatic individuals and in females and males. **(A-C)** Antibody titers over time post-PCR confirmation of asymptomatic subjects for RBD (A), S2 (B), and N (C). **(D-F)** Antibody titers over time post-onset of SARS-CoV-2 infection symptoms from PCR+ confirmed patients or ELISA seropositive PRNT_90_+ individuals from community wide cohort for RBD (A), S2 (B), and N (C), grouped by patient sex. **(G)** PRNT_90_ values over time post-onset of SARS-CoV-2 infection symptoms grouped by patient sex, p value calculated as described previously.

**Table S1.**
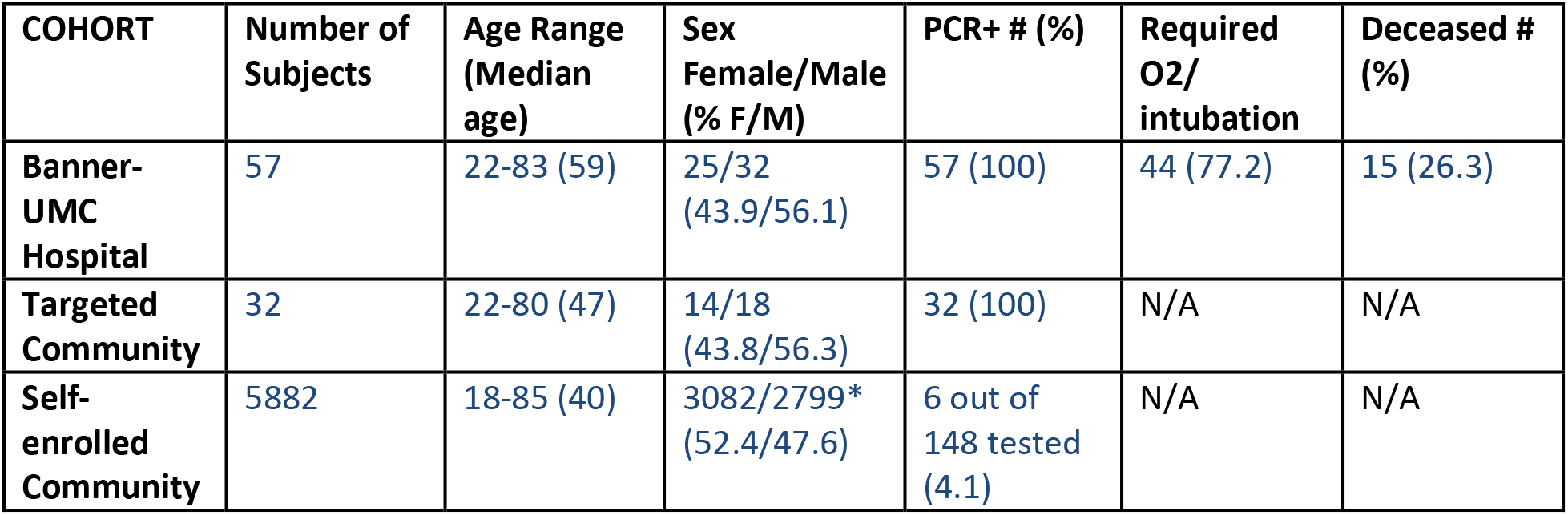
Demographics and essential clinical characteristics of subjects analyzed in this study. Recruitment started in early April, 2020, and included the groups described above and in Methods. Banner-UMC group was restricted to hospitalized subjects; Targeted Community included subjects recruited via fliers and word-of-mouth within faculty, staff and contacts of University of Arizona and Banner-UMC. Self-enrolled community were enrolled via website into the UArizona Antibody Testing Study, supported by the State of Arizona contract; the testing was open to community, first responders and health care workers in Tucson, AZ. *1 intersex participant.

| COHORT | Number of Subjects | Age Range (Median age) | Sex Female/Male (% F/M) | PCR+ # (%) | Required O2/ intubation | Deceased # (%) |
| --- | --- | --- | --- | --- | --- | --- |
| Banner-UMC Hospital | 57 | 22-83 (59) | 25/32 (43.9/56.1) | 57 (100) | 44 (77.2) | 15 (26.3) |
| Targeted Community | 32 | 22-80 (47) | 14/18 (43.8/56.3) | 32 (100) | N/A | N/A |
| Self-enrolled Community | 5882 | 18-85 (40) | 3082/2799* (52.4/47.6) | 6 out of 148 tested (4.1) | N/A | N/A |

## METHODS

### Human subjects

All human subject work was approved by the University of Arizona IRB and was conducted in accordance with all federal, state and local regulations and guidelines under the protocols # 1510182734 and 1410545697A048. Human subject group characteristics are described in Supplementary Table 1, as well as below in the text. Subjects were recruited in three ways. First, targeted recruitment was used to recruit confirmed positive COVID-19 PCR test subjects with severe COVID-19, defined as one that needed hospitalization into the Banner-University Medical Center. Second, targeted recruitment was used to recruit subjects with confirmed positive COVID-19 PCR test who did not require hospitalization (mild/moderate COVID-19 cases). Finally, the vast majority of subjects were recruited via public announcement and website registration as part of the University of Arizona Antibody Testing Pilot. Following website registration, subjects were consented and bled. Blood was centrifuged at six sites across Tucson, AZ, between April 30 and May 7^th^. For all subjects, venous blood was obtained by venipuncture into SST Vacutainer tubes (Becton-Dickinson, Sunnyvale, CA, cat. #367988), serum separated by centrifugation at 1,200 rpm and sent to the central processing laboratory within 4 h. For both hospitalized and non-hospitalized targeted recruitment groups, following aliquoting, serum was used for the ELISA assay with or without freezing and thawing as described below. Finally, sera from 352 subjects recruited into the above two IRB protocols prior to September, 2019, served as negative controls for assay development. Based on local and general prevalence, it would be expected that 96-98% of these subjects have previously encountered seasonal coronaviruses (Gorse et al., 2010). Freezing and thawing had no effect on levels of antibodies detected by ELISA or PRNT.

### Virus

SARS-Related Coronavirus 2, Isolate USA-WA1/2020 (BEI NR-52281) was passaged once on Vero (ATCC #CCL-81) cells at a MOI of 0.01 for 72 hours. Supernatant and cell lysate were combined, subjected to a single freeze-thaw, and then centrifuged at 3000RPM for 10 minutes to remove cell debris.

### Antigens and Antiviral antibody assay

The bacterially-produced recombinant receptor-binding domain (RBD) of the spike (S) glycoprotein was a gift of Dr Daved Fremont (Washington University, St. Louis, MO). Mammalian RBD was purchased from Genscript (catalog # Z03483). SARS-CoV-2 N (nucleocapsid) protein was purchased from Genscript (catalog # Z03488), and S2 subdomain of the SARS-CoV-2 S glycoprotein was purchased from Sino Biological (catalog # 40590-V08B).

Enzyme-linked immunosorbent assay (ELISA) was performed as described (Amanat et al., 2020) with several minor modifications. To obtain titers, antigens were immobilized on high-adsorbency 96-well plates at 5 ng/ml. Plates were blocked with 1% non-fat dehydrated milk extract (Santa Cruz Biotechnology #sc-2325) in sterile PBS (Fisher Scientific Hyclone PBS #SH2035,) for 1 hour, washed with PBS containing 0.05% Tween-20, and overlaid with serial dilutions of the serum or plasma for 60 min. Plates were then washed and incubated for 1hr in 1% PBS and milk containing an anti-human IgG-HRP conjugated antibody (Jackson Immunoresearch catalog 709-035-149) at a concentration of 1:2000 for 1 hour. For IgM detection an anti-human IgM-HRP conjugated antibody (Jackson Immunoresearch catalog 709-035-073) was used at a concentration of 1:5000 and incubated for 1 hour. Plates were washed with PBS-Tween solution followed by PBS wash. To develop, plates were incubated in tetramethylbenzidine prior to quenching with 2N H2SO4. Plates were then read for 450nm absorbance.

ELISAs on community-wide samples were performed at the University of Arizona Genomics Core. A 384 well format was applied for high throughput screening, with protocol conditions remaining identical except for the substitution of anti-human Pan-Ig HRP conjugated antibody (Jackson Immunoresearch catalog 109-035-064). Plates were read for 450nm absorbance on CLARIOstar Plus from BMG Labtech. Samples with OD630 values greater than 0.05 were re-run. Every plate contained at least 32 seronegative controls and either CR3022 or HM3128 (Creative Diagnostics) monoclonal antibodies as a positive control for RBD or S2, respectively.

### Plaque reduction neutralization test

A plaque reduction neutralization test (PRNT) for SARS-CoV-2 was developed based on our prior work (Uhrlaub et al., 2011). Briefly, Vero cells (ATCC # CCL-81) were plated in 96 well tissue culture plates and grown overnight. Serial dilutions of plasma/serum samples were incubated with 100 plaque forming units of SARS-CoV-2 for 1 hour at 37° C. Plasma/serum dilutions plus virus were transferred to the cell plates and incubated for 2 hours at 37°C, 5% CO2 then overlayed with 1% methylcellulose. After 72 hours, plates were fixed with 10% Neutral Buffered Formalin for 30 minutes and stained with 1% crystal violet. Plaques were imaged using an ImmunoSpot Versa (Cellular Technology Limited, Cleveland, OH) plate reader. The serum/plasma dilution that contained 10 or less plaques was designated as the NT90 titer.

### Statistical analysis

Statistical analyses are described in the corresponding Figure Legends.

